# Online Personal Training in Patients with Marfan Syndrome: A Randomized Controlled Study of its Impact on Quality of Life and Physical Capacity

**DOI:** 10.1101/2023.05.16.23289922

**Authors:** Steeve Jouini, Olivier Milleron, Ludivine Eliahou, Guillaume Jondeau, Damien Vitiello

## Abstract

**Background:** Marfan syndrome (MFS) is a rare genetic disorder affecting the vascular and musculoskeletal systems. Exercise is classically contraindicated and there are no data on the limitations associated with the syndrome and the benefits of training in this population. This study aimed to characterise the quality of life (QoL) and physical capacity of patients with MFS and to evaluate the benefits of a 3-month online personal training program.

**Methods:** MFS patients were compared with healthy subjects (H-S) at baseline. They were then randomized 1:1 into a training group (MFS-T) and a control group (MFS-C). The training consisted of 2 supervised online training sessions per week at home for 3 months, and the session program was selected based on the initial assessment. The main outcome measure was QoL as assessed by the MOS SF-36. The evolution of parameters during training was compared between MFS-T and MFS-C.

**Results:** At baseline, QoL in all dimensions was lower in MFS. Peak oxygen uptake (VDO_2_peak) was also 25% lower, as was muscle elasticity. Training significantly improved 1) QoL (+20.2±14.3 MFS-T *vs.* 0.7±0.5 MFS-C), 2) VDO_2_peak (+34% MFS-T *vs.* 14% MFS-C), 3) muscle elasticity index (11.5±8.2 MFS-T *vs.* +1.2±1. 7 MFS-C), reduced blood pressures during isometric squat (systolic −19±30 MFS-T *vs.* 0±6 MFS-C; diastolic −27±39 MFS-T *vs.* +2±15 MFS-C), reduced pulse wave velocity (PWV) at rest (-1.20±1.89 MFS-T *vs.* −0.40±1.61 MFS-C) and after peak exercise (-0.42±0.45 MFS-T *vs.* 0.08±0.48 MFS-C). Aorta diameter remained stable in both groups (MFS-T −0.19±1.1 *vs.* 0.11±0.78 MFS-C). After training, QoL remained lower in MFS-T than in H-S, but peak VDO_2_, PWV at rest and after exercise were similar to those of H-S.

**Conclusions:** A 3-month online training program had a beneficial effect on QoL, cardiovascular and muscular parameters in MFS without affecting aortic root diameter.

## Introduction

Marfan syndrome (MFS) is an autosomal dominant genetic disorder caused by a pathogenic variant in the gene encoding fibrillin 1 (*FBN1*), with more than 1,300 unique pathogenic variants reported. MFS affects multiple systems, including the cardiovascular and musculoskeletal systems ^1–4^ and is associated with altered quality of life (QoL) ^5^. The fatal risk associated with MFS is related to aortic aneurysm expansion leading to aortic dissection which can be prevented by prophylactic surgery. To date, the main medical therapy to reduce the cardiovascular impact of MFS is the use of betablockers and regular medical follow-up ^6^. There is currently no specific exercise program available and validated, and recommendations still limit physical activity to minimize the risk of aortic dilation, dissection, and possible aortic rupture ^6^.

The benefits of cardiopulmonary exercise training are well documented in the general population and its importance is increasingly being emphasised in patients with various cardiovascular diseases, including chronic coronary heart disease^7, 8^ and heart failure^9^. However, there is a lack of clear data in patients with MFS. In fact, patients with MFS may differ from other populations because MFS has been associated with a specific myopathy^10^ and there is concern that increasing the blood pressure (BP) during exercise may increase aortic root dilatation and the risk of aortic dissection. Indeed, aortic dissection has been reported following isometric exercise. Deconditioning may result from this concern and may also contribute to the alteration in exercise capacity and QoL in patients with MFS.

In addition, recent animal studies suggest that regular endurance exercise may be beneficial ^11, 12^. Therefore, a personalised online training program in combination with standard care may be beneficial in patients with MFS^13^.

To our knowledge, there are currently no randomized and controlled trials evaluating the effects of such a program in patients with MFS and no data on the physical and physiological capacity of these patients. Therefore, the aim of this study was to evaluate the effects of a 3-month personalised e-training program on QoL, cardiovascular and muscular capacity in adult patients with MFS. The results of this study may provide new insights into the management of MFS patients and contribute to the development of more effective treatment strategies.

## Method

### Study population

MFS patients: Inclusion criteria for MFS patients were: 1) adult patients (18 to 75 years of age); 2) presence of an FBN1 pathogenic variant; 3) ability to exercise; and 4) having health insurance. Exclusion criteria for MFS patients were: patients with cardiovascular disease unrelated to MFS, pregnant patients, history of aortic dissection, aortic diameter > 45 mm, significant aortic regurgitation, uncontrolled resting hypertension (diastolic blood pressure > 90 mmHg and systolic blood pressure > 140 mmHg), unavailability by telephone, participation in an experiment in the 3 months prior to screening, unwillingness or inability to sign the informed consent form.

Healthy subjects were matched for age and sex.

### Study Design

MFS patients were randomized 1/1 into 2 groups: 1) MFS-C who did not benefit from the training program and were assessed at baseline and after 3 months; 2) MFS-T who followed the personalised e-training program for 3 months and were assessed at baseline and after 3 months of training.

All participants gave written informed consent before enrolment. The study complied with the Declaration of Helsinki, the institutional ethics committee approved the study protocol (#2020-A01751-38) and the French Society of Cardiology promoted the study.

It has been approved by Protection of Persons Committee SOUTH MEDITERRANEAN CHU CIMIEZ HOSPITAL CS 91179 06003 NICE CEDEX 1.

The trial was prospectively registered at Clinical Trial NCT04553094

## Evaluation of patients

### Quality of life assessment

The Medical Outcome Study Short Form - 36 (MOS SF-36) was used to assess QoL.^14^.

### Echocardiography

Patients with MFS underwent standard echocardiography and tissue Doppler imaging (Vivid 9 Dimension® ultrasound device - GE Healthcare). This study included calculation of left ventricular ejection fraction (LVEF), calculation of E/A ratio. Aortic diameters were measured at different levels (*i.e.,* ring, root, tubular aorta, arch, descending thoracic aorta and abdominal aorta). 2D strain echocardiography was also performed to assess global systolic longitudinal strain (GLS) of the LV and right ventricle (RV).

### Body composition

Body composition was measured using a bioimpedance scale (Tanita Body Composition Analyzer BC-420MA). Body weight and percentage of fat mass (BF%), muscle mass (MM%) and water were assessed. Body mass index (BMI) was calculated using the following formula BMI = kg / m2, where kg is the body weight in kilograms and m2 is the height in metres squared.

### Cardiopulmonary exercise training (CPET) and spirometry

At rest, before exercise, an electrocardiogram (ECG) was taken (200S-Cardioline), SBP and DBP were measured with an automatic tensiometer (METRONIK BL-6 1000) and pulse wave velocity (PWV) was measured with a Popmètre®.

The patients then performed an incremental exercise on the ergocycle to assess their cardiorespiratory capacity. The intensity of the exercise gradually increased by 10 W/min until the patients voluntarily stopped. During the exercise, the patients’ expired gases were continuously analysed. Peak oxygen uptake (VDO2peak) was determined as the highest value achieved during exercise. The ventilatory threshold 1 (VT1), the oxygen uptake efficiency slope (OUES), the ratio of ventilation (VE) to carbon dioxide production (VDCO_2_) (VE/ VDCO_2_) were calculated. Pulse wave velocity (PWV) was measured after the first ventilator threshold.Heart rate (HR) and SBP And DBP were measured every 3 minutes, during the CPET. If the SBP > 160 mmHg during the exercise (*i.e.,* CPET), the patient with MFS was excluded from the study ^15–18^.

Pulse wave velocity (PWV) was measured again after exercise.

HR at the first ventilatory threshold (VT1) (HRVT1) and HR at VDO_2_peak (HRpeak VDO_2_) were recorded to adapt and personalise the training loads for the 3-month e-training program. Peak and training HR were determined during CPET.

Forced vital capacity (FVC), forced expiratory volume in one second (FEV1) and peak expiratory flow 25-75% (PEF25-75) were measured and FEV1/FVC was calculated.

### Muscular exercise testing (mET)

Lower limb muscle strength and maximal force contraction were measured during a countermovement jump (cm), a squat jump (cm) and a one repetition maximum (1RM) based on 3 consecutive squats using a linear encoder (Bosco System Platform Chronojump).

They then performed an isometric bodyweight squat. During the squat, blood pressure and PWV were measured (Detailed Appendix 1).

Two vertical jump tests were performed: the countermovement jump (CMJ) and the squat jump (SJ) using the Chronojump platform. The recovery value is the difference measured between the heights of the two jumps.

### Pulse wave analysis

Pulse wave analysis was performed at rest, during exercise (*i.e.,* isometric squat and CPET) and after exercise. Complete pulse wave analysis included cardiac index, reflection coefficient, PWV, AIx, SBP and DBP.

- At rest before the exercise test. A comprehensive analysis was first performed at rest in the supine position. The average of the three consecutive results obtained was used for analysis.
- During the squat. Pulse wave velocity (PWV), systolic blood pressure (SBP) and diastolic blood pressure (DBP) were then measured during the ’isometric squat’ while the patients were squatting with their knees bent at 90°, positioned on a step with their toes halfway over the edge of the step. One sensor was placed on the second toe and the other on the middle finger of the hand, while the cuff was placed on the left arm. Only one assessment of arterial pressure was performed and PWV was measured (PWV was continuously monitored).
- At rest after the CPET exercise test. Finally, the complete analysis was repeated at rest in the supine position after the CPET (average of 3 measurements).

### 3-month personalized online training program for Marfan patients

The MFS patients in the MFS-T group completed the personalised 3-month e-training program at home. The program consisted of 2 training sessions per week for a total of 24 sessions. The initial exercise intensity during the training sessions was chosen based on HRVT1 and the HR peak achieved during CPET. The patient had to remain within this intensity range.

Details of the training method and evaluations are described in Appendices 1 and 2. HR, DBP and SBP were telemonitored during each training session and followed live either by videoconference and/or by connected devices. This made it possible to check that patients were achieving the target HR and to verify that SBP did not exceed 160 mmHg. Training loads were adjusted weekly to achieve the best training effect based on heart rate and the RPE (rate of perceived exertion) scale ^19, 20^. Finally, patients were frequently reminded of the training program, 3 to 4 training sessions in advance, in order to increase their adherence to the program.

### Statistical analyses

The study used 1:1 randomisation to form treatment and control groups to minimise potential selection bias and maximise statistical power. The sample size was determined based on the study objectives using 80% power and a 5% significance level. A mean difference of 13 points in QoL (physical component) with a standard deviation of 20 was estimated between the trained group and the control group. The minimum number of subjects required in each group was 38, but to allow for a 20% drop-out rate, the number of patients in each group was increased to 46.

Statistical analyses were performed using GraphPad Prism 9.4.1 software (San Diego, CA, USA). One-way ANOVA was used to compare the different groups (H-S, MFS-C, MFS-T) in the pre-and post-training sessions. Two-way repeated measures ANOVAs were used to identify pre-post differences in the MFS-C and MFS-T groups. Different models (*i.e.,* time, group and group x time interaction) were used. When significant interactions were found, Tukey’s post hoc test was used. To compare the effect of training on the MFS-C and MFS-T groups, we analysed the difference after 3 months - baseline and used a Student’s t-test. Data were presented as percentages (%) or mean ± standard deviation. Statistical significance was considered when P values were ≤ 0.05.

## Results

A total of 105 subjects were recruited for the study (Figure 1). There were 70 MFS patients and 35 healthy subjects. The characteristics of the participants are shown in Table 1. The H-S were sex and age matched to the MFS patients, all of whom were receiving beta-blockers.

**Figure: 1.**
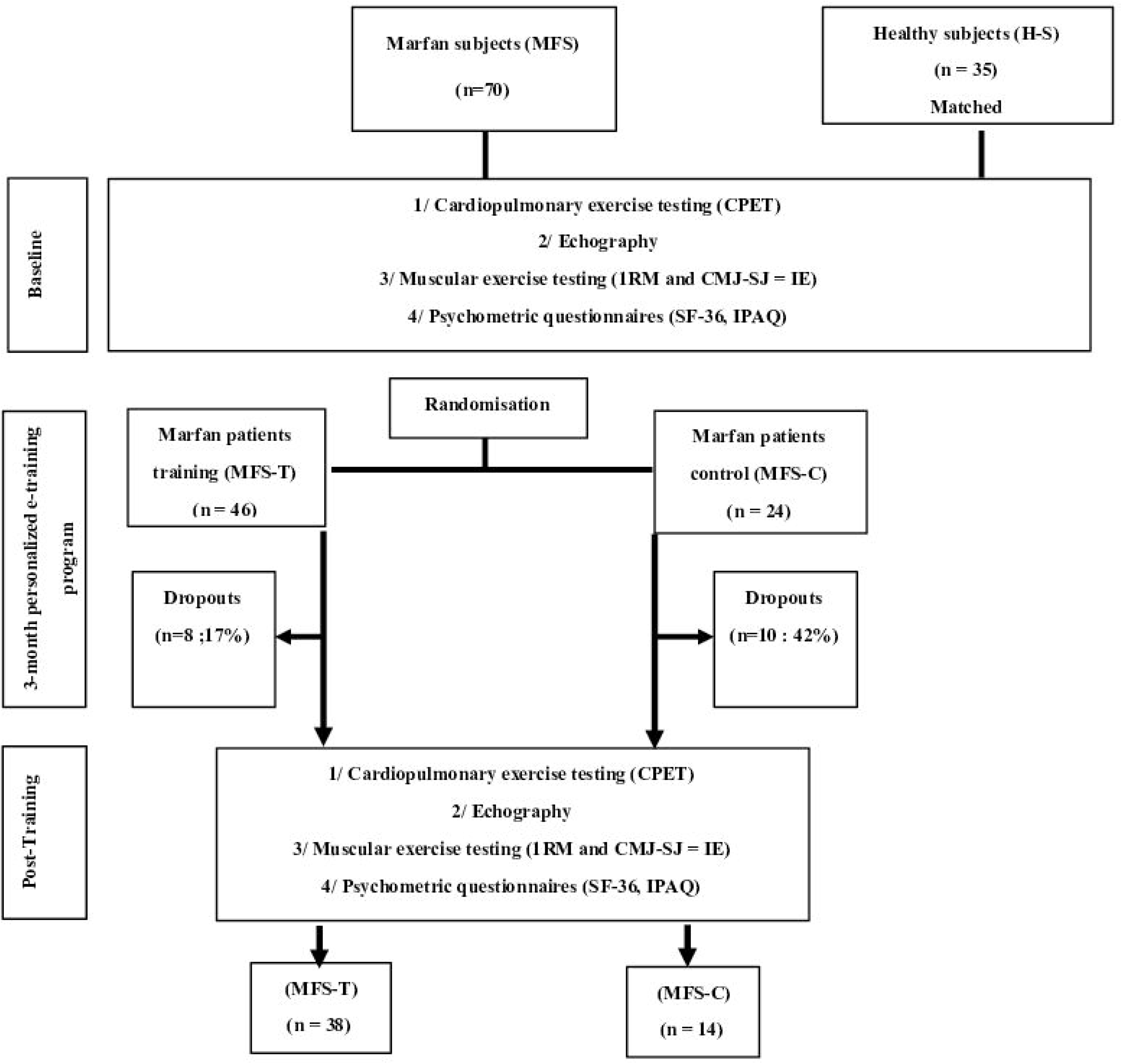
Flow chart.

**Table 1:**
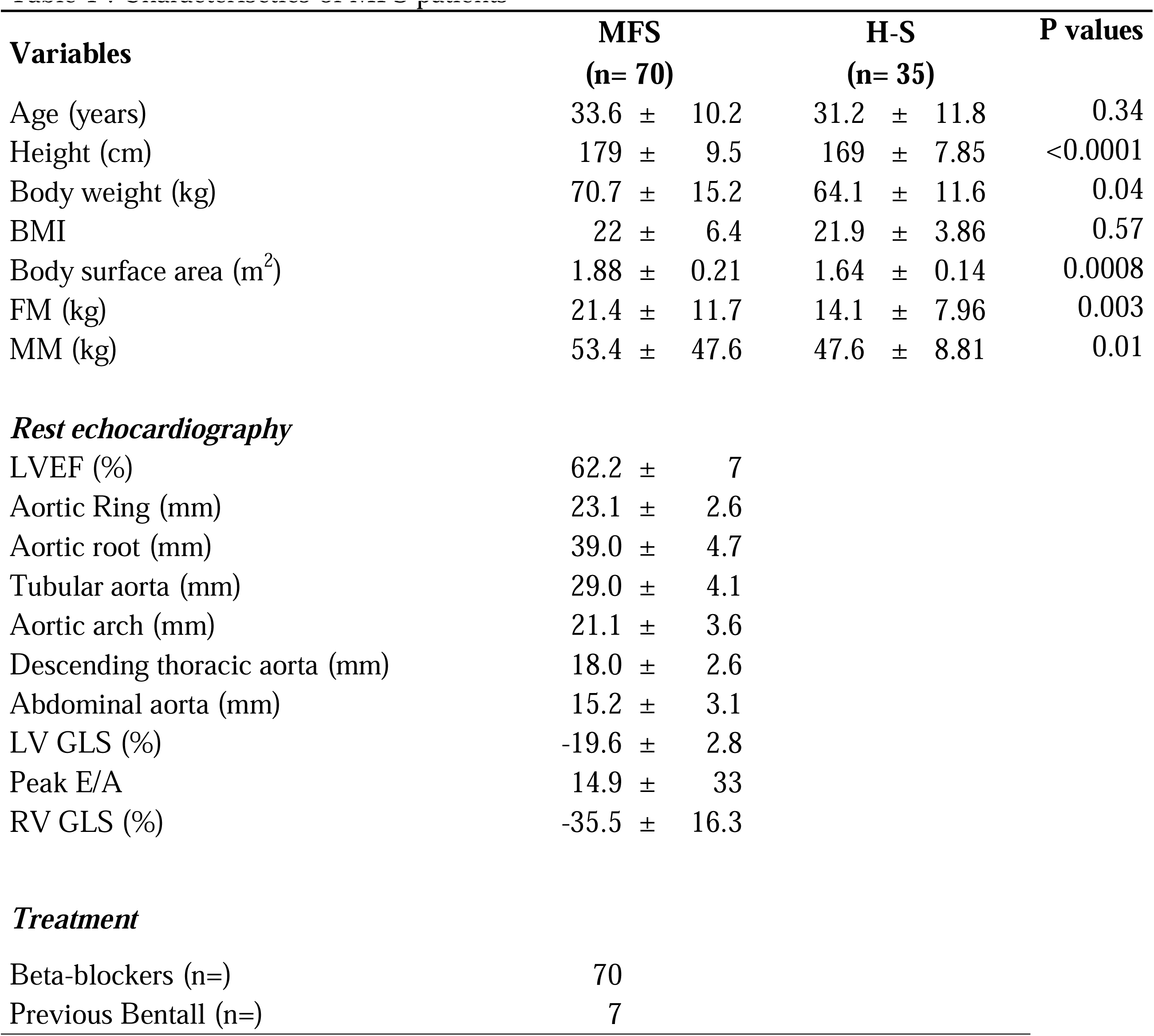
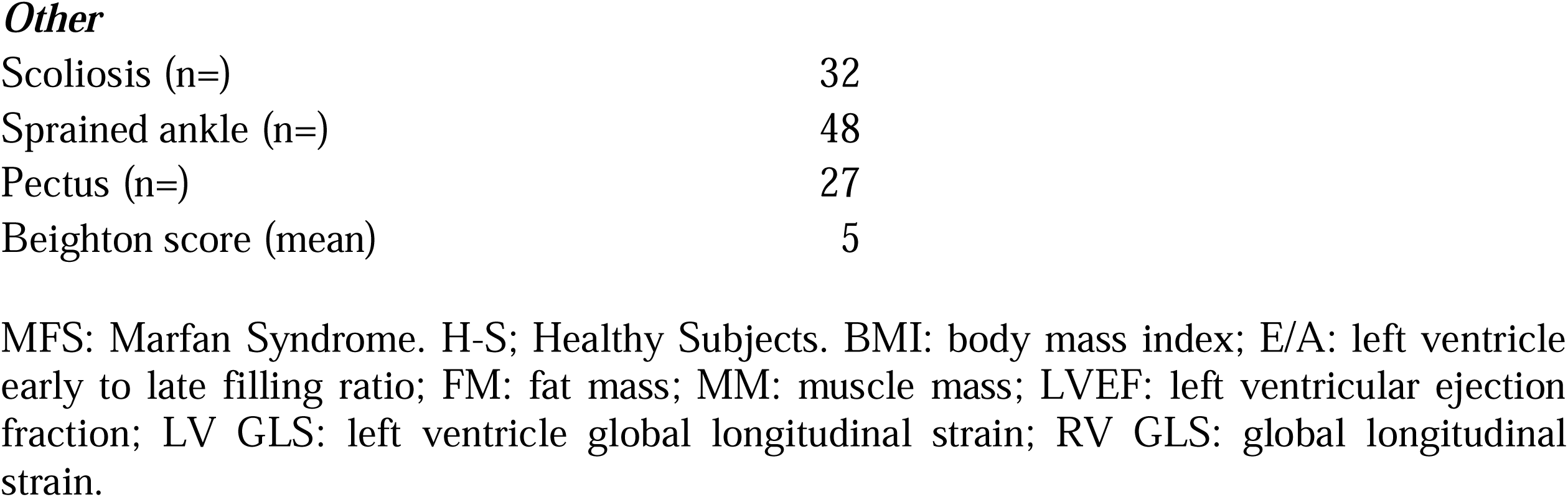
Characterisctics of MFS patients

| Variables | MFS<br>(n= 70) |  | H-S<br>(n= 35) |  | P values |
| --- | --- | --- | --- | --- | --- |
| Age (years) | 33.6 | ± 10.2 | 31.2 | ± 11.8 | 0.34 |
| Height (cm) | 179 | ± 9.5 | 169 | ± 7.85 | <0.0001 |
| Body weight (kg) | 70.7 | ± 15.2 | 64.1 | ± 11.6 | 0.04 |
| BMI | 22 | ± 6.4 | 21.9 | ± 3.86 | 0.57 |
| Body surface area (m<sup>2</sup>) | 1.88 | ± 0.21 | 1.64 | ± 0.14 | 0.0008 |
| FM (kg) | 21.4 | ± 11.7 | 14.1 | ± 7.96 | 0.003 |
| MM (kg) | 53.4 | ± 47.6 | 47.6 | ± 8.81 | 0.01 |
| ***Rest echocardiography*** |  |  |  |  |  |
| LVEF (%) | 62.2 | ± 7 |  |  |  |
| Aortic Ring (mm) | 23.1 | ± 2.6 |  |  |  |
| Aortic root (mm) | 39.0 | ± 4.7 |  |  |  |
| Tubular aorta (mm) | 29.0 | ± 4.1 |  |  |  |
| Aortic arch (mm) | 21.1 | ± 3.6 |  |  |  |
| Descending thoracic aorta (mm) | 18.0 | ± 2.6 |  |  |  |
| Abdominal aorta (mm) | 15.2 | ± 3.1 |  |  |  |
| LV GLS (%) | -19.6 | ± 2.8 |  |  |  |
| Peak E/A | 14.9 | ± 33 |  |  |  |
| RV GLS (%) | -35.5 | ± 16.3 |  |  |  |
| ***Treatment*** |  |  |  |  |  |
| Beta-blockers (n=) | 70 |  |  |  |  |
| Previous Bentall (n=) | 7 |  |  |  |  |

***Other***
|  |  |
| --- | --- |
| Scoliosis (n=) | 32 |
| Sprained ankle (n=) | 48 |
| Pectus (n=) | 27 |
| Beighton score (mean) | 5 |

MFS: Marfan Syndrome. H-S; Healthy Subjects. BMI: body mass index; E/A: left ventricle early to late filling ratio; FM: fat mass; MM: muscle mass; LVEF: left ventricular ejection fraction; LV GLS: left ventricle global longitudinal strain; RV GLS: global longitudinal strain.

The MFS patients were heavier and taller than the H-S. Almost half of the MFS patients had scoliosis and 61% had ankle sprains. The MFS patients also had dilation of the aorta and global joint hypermobility.

### Baseline : Comparison of MFS patients with matched controls

QoL (Table 2) was significantly different between MFS and H-S in all eight dimensions assessed (P < 0.05). Physical function, the criterion used to calculate the power of the study, was 55% and 68.97% lower in MFS-C and MFS-T, respectively, compared to H-S.

**Table 2:**
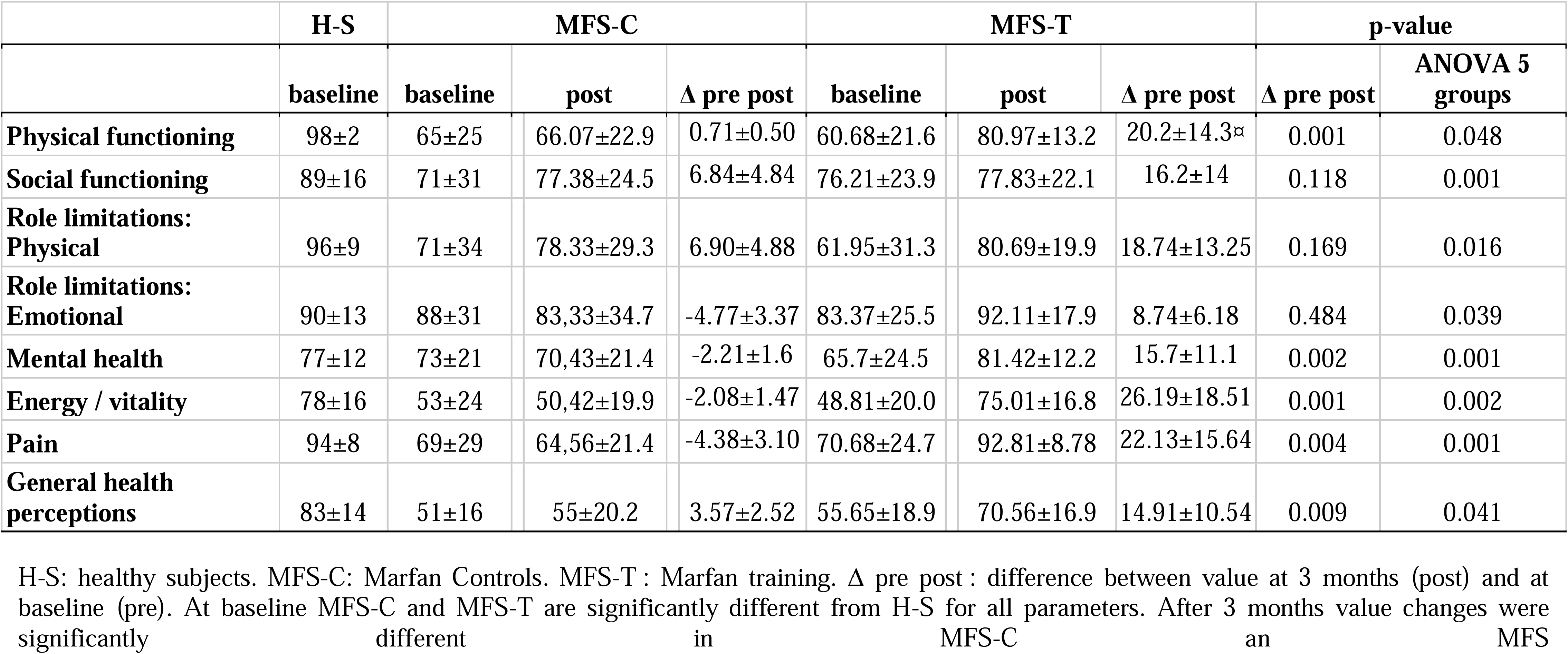
The quality of life and its evolution with training in MFS patients

VDO_2_peak was 25% lower in MFS (MFS-C and MFS-T) compared to H-S (Figure 2a). PWV was significantly higher in MFS immediately after CPET (Figure 2b), (P<0.001). SBP and DBP were significantly higher in MFS than in H-S during isometric squat (+13% and +44% respectively; Figure 2c).

**Figure 2a:**
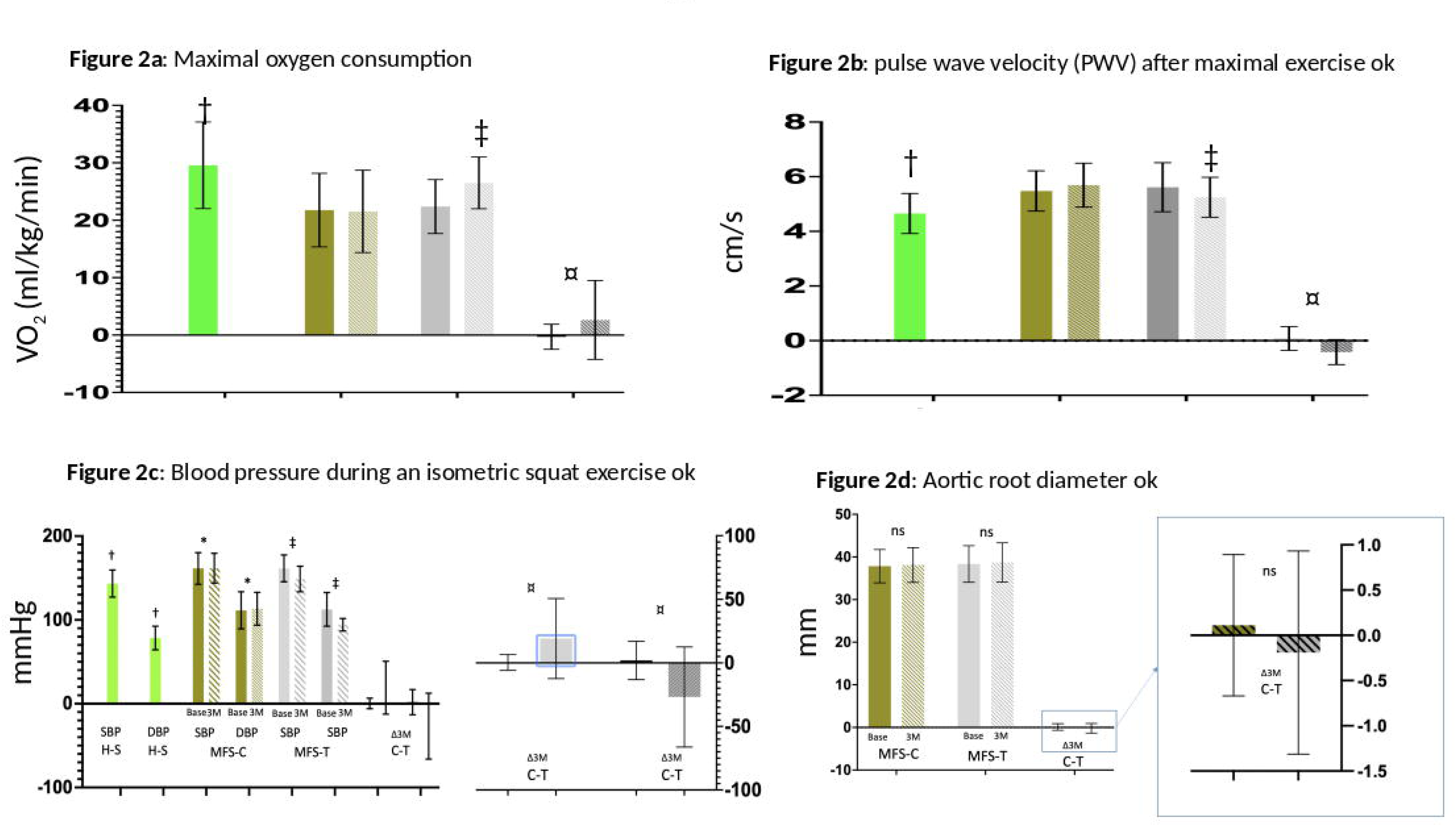
Peak oxygen consumption. Peak oxygen consumption (VLO_2_peak) in H-S (healthy subjects), MFS-C (Marfan syndrome control), and MFS-T (Marfan syndrome training), at baseline (base) and after 3 months (3M). Δ3M is the difference after 3 months compared to baseline. † p<0,05 when compared to MFS-C Base and MFS-T Base. ‡ p<0,05 *vs.* MFS-T Base and MFS-C Base and post. ¤: The Δ3M is significantly greater in MFS-T than in MFS-C (p<0,05).

**Figure 2b.** Pulse wave velocity. 2b: Pulse wave velocity (PWV) after peak exercise. in H-S (healthy subjects), MFS-C (Marfan syndrome control), and MFS-T (Marfan syndrome training), at baseline (base) and after 3 months (3M). Δ3M is the difference after 3 months compared to baseline. † p<0,05 when compared to MFS-C Base and MFS-T Base. ‡ p<0,05 *vs.* MFS-T Base and MFS-C Base and post. ¤: The Δ3M is significantly greater in MFS-T than in MFS-C (p<0,05).

**Figure 2c.** Blood pressure during an isometric squat exercise. Blood pressure (BP = SBP and DBP) is measured during an isometric squat exercise in H-S (healthy subjects), MFS-C (Marfan syndrome control), and MFS-T (Marfan syndrome training), at baseline (base) and after 3 months (3M). Δ3M is the difference after 3 months compared to baseline. † p<0,05 when compared to MFS-C Base and MFS-T Base. ‡ p<0,05 *vs.* MFS-T Base and MFS-C Base and post. ¤: The Δ3M is significantly greater in MFS-T than in MFS-C (p<0,05).

**Figure 2d.** Aortic root diameter Aortic root diameter is measured during an isometric squat exercise MFS-C (Marfan syndrome control), and MFS-T (Marfan syndrome training), at baseline (base) and after 3 months (3M). Δ3M is the difference after 3 months compared to baseline. No significant diffence were observed

The 1RM values were significantly lower in MFS patients compared to H-S (p < 0.001) (Figure 3a).

**Figure 3a:**
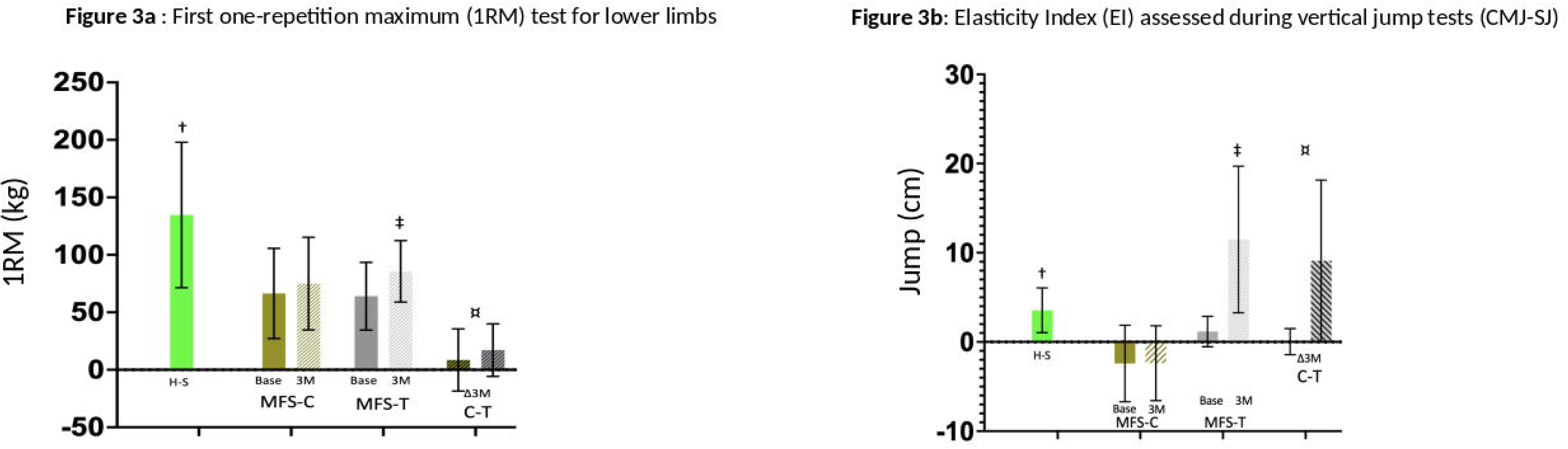
One repetition maximum (1RM) test for lower limbs. time of the one repetition maximum (1RM). in H-S (healthy subjects), MFS-C (Marfan syndrome control), and MFS-T (Marfan syndrome training), at baseline (base) and after 3 months (3M). Δ3M is the difference after 3 months compared to baseline. † p<0,05 when compared to MFS-C Base and MFS-T Base. ‡ p<0,05 *vs.* MFS-T Base and MFS-C Base and post. ¤: The Δ3M is significantly greater in MFS-T than in MFS-C (p<0,05).

**Figure 3b:** Elasticity Index (EI) assessed during vertical jump tests (CMJ-SJ) the elasticity index (EI) was assessed during vertical jump tests (CMJ-SJ) . in H-S (healthy subjects), MFS-C (Marfan syndrome control), and MFS-T (Marfan syndrome training), at baseline (base) and after 3 months (3M). Δ3M is the difference after 3 months compared to baseline. † p<0,05 when compared to MFS-C Base and MFS-T Base. ‡ p<0,05 *vs.* MFS-T Base and MFS-C Base and post. ¤: The Δ3M is significantly greater in MFS-T than in MFS-C (p<0,05).

The aortic root diameter was similar in the MFS-C and MFS-T groups (38.1±4.1 mm *vs*. 38.7±4.6 mm, NS) and did not increase after training (Figure 2d).

### Characteristics of training sessions

The duration of the training session, peak and mean heart rate during the training session, perceived exertion during the training session and blood pressure at the beginning, middle and end of the training session were recorded and are shown in Table 3.

**Table 3 :** Key parameters during training sessions for MFS patients.

|  | **Duration**<br>**of training** | **HR peak** | **HR mean** | **RPE** | **BP1** | **BP2** | **BP3** |
| --- | --- | --- | --- | --- | --- | --- | --- |
| **Mean** | 987.5 | 145.2 | 116.8 | 6.41 | 123.1/76 | 132.2/78.7 | 126.3/74.9 |
| **SD** | 297.7 | 15.08 | 10.71 | 0.89 | 9.66/5.16 | 9.52/6.75 | 10.91/6.11 |

Duration of training: total training time during protocol. HR peak: peak heart rate reached during training sessions. HR means Mean heart rate during the entire training sessions. RPE: Rated Perceived Exertion Scale measured at the end of training session. BP1 Blood pressure before starting the training session. BP2: blood pressure at mid-session, BP3: blood pressure at the end of training session.

### Main effects of a 3-month personalized online training program in MFS patients: ***Δ***: post - pre

All dimensions of QOL improved significantly more in the MFS-T group than in the MFS-C group after training (Table 2). The main criterion, physical function, increased by 33% in the MFS-T group but remained significantly lower than in the H-S group (P < 0.0001). In addition, a significant 31% reduction in pain was observed in the MFS-T group (P < 0.0001). V O_2peak_ increased significantly more in the MFS-T group that in the MFS-C group (P<0.05) (Figure 2a). Actually, after training, V O_2peak_ in MFS-T group was not significantly different from V O_2peak_ in H-S (P=0.057). (26.5±4.8 ml.min^-1^.kg^-1^ *vs.* 29.4±7.6 ml.min^-1^.kg^-1^, NS). PWV immediately after CPET decreased significantly more in the MFS-T group than in the MFS-C group (5.24±0.9 m/s-1 *vs.* 5.69±0.74 m/s-1, P<0.0001) (Figure 2b). In fact, the post-training PWV in the MFS-T group was close to the PWV in the H-S group (5.24±0.74 m.s-1 MFS-T *vs.* 4.65±0.73 m.s-1 H-S, NS).

1 RM increased significantly more in MFS-T than in MFS-C (+17 kg *vs.*+ 9 kg, P<0.001) (Figure 3a) although it remained significantly lower than in H-S (81.9±21.9 kg *vs.* 133±77 kg, P<0.05). The Elasticity Index increased significantly more in the MFS-T than in the MFS-C (+9.09±1.46 cm MFS-T *vs.* 0.03±1.45 cm MFS-C, P < 0.001) (Figure 3b).

Blood pressure decreased significantly more in the MFS-T than in the MFS-C during isometric squat exercise (sBP −19±30 MFS-T *vs.* 0±6 MFS-C; dBP −27±39 MFS-T *vs.* +2±15 MFS-C; P < 0.02). SBP and dBP were lower after training in the MFS-T group than in the MFS-C group (sBP: 148±15 mmHg MFS-T *vs.* 162±18 mmHg MFS-C, P < 0.05; dBP: 94±8 mmHg±18 mmHg MFS-T *vs.* 113±20 mmHg MFS-C, P < 0.001) (Figure 2c). In the MFS-T group, sBP after training was similar to that in the H-S group (148±11 mmHg *vs.* 143±16 mmHg, NS), but dBP tended to be higher (94±8 mmHg±7.37 mmHg *vs.* 78±14 mmHg, NS). Aortic root diameters remained stable and were similar in MFS-T and MFS-C after training (38.1±4.1 mm *vs.* 39.1±4.4 mm, NS; Figure 2d). In fact, during training, aortic diameter tended to decrease in MFS-T and increase in MFS-C, but this difference was not statistically significant (0.11±0.78 *vs.* −0.19±1.12 mm; p=0.74) (Figure 2d).

### Cardiovascular and respiratory effects of a 3-month personalized e-training program in MFS patients

The e-training program reduced the Aix and the reflection coefficient and increased the elasticity index in the MFS-T group compared to the MFS-C group (Table 4). It also reduced PWV at rest, during a squat and after CPET in the MFS-T group compared to the MFS-C group. The reduction in SBP with training was greater in the MFS-T group compared to the MFS-C group at rest, during a squat and during the recovery period immediately after CPET, but not during CPET. Finally, no significant differences in Tiffeneau index and PEF 25-75 were found between MFS-C and MFS-T after the exercise program.

**Table 4:** Cardiovascular and pulmonary effects of training in MFS patients.

| Variables ($\Delta$ pre-post) | MFS-C | | | MFS-T | | | P values |
| --- | --- | --- | --- | --- | --- | --- | --- |
| LVEF (%) | -0.14 | ± | 8.65 | 5.24 | ± | 15.4 | 0.391 |
| Cardiac Index (L/min/m<sup>2</sup>) | 0.05 | ± | 0.83 | 0.05 | ± | 0.92 | 0.990 |
| Reflection coefficient (%) | -0.30 | ± | 10.7 | -12.4 | ± | 14.5 | 0.020 |
| Alx (%) | -3.10 | ± | 12.9 | -12.2 | ± | 9.5 | 0.024 |
| EI (cm) | 0.13 | ± | 1.52 | 1.89 | ± | 1.99 | 0.009 |
| PWV at rest (m/s) | -0.02 | ± | 0.31 | -0.45 | ± | 0.45 | 0.011 |
| PWV during squat (m/s) | 0.60 | ± | 1.02 | -2.99 | ± | 2.12 | 0.001 |
| PWV post CPET (m/s) | 0.95 | ± | 0.75 | -0.87 | ± | 1.69 | 0.001 |
| Systolic BP at rest (mmHg) | 1.46 | ± | 5.11 | -2.20 | ± | 6.13 | 0.070 |
| Systolic BP during squat (mmHg) | 3.50 | ± | 6.42 | -13.5 | ± | 16.9 | 0.008 |
| Systolic BP during CPET (mmHg) | 12.9 | ± | 30.7 | -0.57 | ± | 21.8 | 0.192 |
| Systolic BP post CPET (mmHg) | 5.36 | ± | 4.89 | -3.65 | ± | 5.46 | 0.001 |
| FEV1/FVC (%) | 0.91 | ± | 16.8 | 4.30 | ± | 7.33 | 0.224 |
| PEF 75-25 (%) | 0.19 | ± | 0.80 | 4.44 | ± | 18.10 | 0.445 |

$\Delta$ : pre to post variation. Alx: augmentation index. BP: blood pressure. EI: Elasticity Index.
LVEF: left ventricular ejection fraction. MFS-C: Marfan control. MFS-T: Marfan training. PEF:
Peak expiratory flow. PWV: pulse wave velocity. FEV1/FVC%: Tiffeneau index.

### Dropout rates

The overall dropout rate was 25%. However, it was lower in the MFS-T group (17%) than in the MFS-C group (42%) (Figure 1).

## Discussion

In this randomized and controlled study, we show that 3 months of personalised home-based training significantly restores the QoL of MFS patients, in particular the main criterion "physical function", increases exercise capacity as measured by VDO_2_peak and muscle strength as measured by 1RM, and improves arterial compliance as shown by a decrease in PWV. These changes can be attributed to the training program we proposed to our patient, thanks to the randomized control design of our study, which made it possible to compare the evolution of each parameter after the 3-month period in the 2 groups (control group, without training, called MFS-C, and training group, called MFS-T). The results indicate a beneficial cardiopulmonary, muscular and vascular effect of training in this population. In addition, training reduced resting and exercise sBP and dBP, which may also be beneficial in the long term, and no detrimental effect on aortic root diameter was observed.

In our program, each exercise session was individualised based on the CEPT at baseline. The intensity of the exercise was chosen to keep the heart rate above the heart rate observed at the first ventilator threshold during CPET and below the peak heart rate observed during CPET. The first training session was conducted in the hospital under medical supervision to monitor blood pressure and heart rate during the exercises proposed for the training sessions. All subsequent sessions were carried out at home. During the course of the program, the intensity of the home training sessions was adjusted on the basis of the perceived exertion scale and the heart rate recorded by the patient during the previous training session. This allowed for a personalised home-based program with maximum safety and efficiency.

To our knowledge, there is only one study of exercise training in people with MFS ^15^. In this observational study of a 3-week low-intensity exercise program in 18 MFS patients, the authors reported positive effects on mental health, fatigue and exercise capacity. The effect on aortic root diameter was not reported in this paper. However, a beneficial effect on the aortic root has been suggested in MFS mice^11, 12^ : in this mouse model, moderate-intensity exercise reduced the growth rate of aortic diameter and the risk of aortic rupture in exercised mice compared with their sedentary counterparts. These protective effects were achieved with exercise at intensities between 55 and 65% of VDO_2_peak.

Changes in QoL in patients with Marfan syndrome have been reported previously^21, 22^. However, no intervention has been shown to improve QoL in this population. The beneficial effects of exercise training on QoL in patients with cardiovascular disease have been well documented ^23, 24^, but data on the effects of training in patients with MFS are scarce. We show that exercise improves all dimensions of QoL (Table 4). The dropout rate may also indirectly reflect the perceived benefit of the training program by patients: in our study, the dropout rate was much higher in the control group than in the training group (42% *vs.* 17%). Training may become an important tool to limit the decline in QoL previously reported in the MFS population^22^.

The functional limitation of MFS patients is evidenced by the 25% lower V O_2peak._at baseline when compared to healthy subjects. This result is in keeping with two previous small studies ^25, 26^, and may partly explain the altered QoL in this population. Betablockade may participate in this decreased peak VDO_2_, as all MFS patients were receiving this therapy: in normal suhjects^27^, betablockade has been associated with decreased peak VDO_2_ by 5 to 15%, and therefore it is unlikely that betablockade does account for the entire difference between healthy subjects and MFS patients. Beyond VDO_2_, a number of parameters are altered in this population, such as maximal strength and elasticity index in the muscular component of the lower limbs (already reported by 2 previous studies ^25, 26^) which may also participate in the altered QoL.

The training program may be beneficial beyond the increase in peak VDO_2_ and improved QoL: the training program increased pulmonary parameters such as Tiffeneau index and PEF25-75%, which increased to values similar to those of healthy subjects (Table 3). The 3 muscle components were also increased: 1/ increase in muscle mass, 2/ increase in muscle strength, 3/ improvement in muscle elasticity. Finally, and perhaps more importantly, PWV was increased at baseline, during CPET and during the recovery phase, as was the augmentation index (Aix), all of which indicate more rigid arteries in the patients. Increased stiffness may be responsible for greater recoil and therefore increased stress on the proximal aorta, which has been associated with increased aortic root dilatation (ref) and may be related to the altered vasodilator mechanism in response to acute exercise in MFS patients ^6, 28^. During the exercise program, PWV and Aix decreased. These effects may be beneficial in the long term and are consistent with results from animal studies showing that exercise improves aortic cellular structure and, in particular, arterial compliance ^11, 12^. This beneficial effect should be additive to the expected benefit associated with lower blood pressure resulting from the lowering effect of training on blood pressure rise during exercise. Because of all these indirect haemodynamic effects, one would expect that exercise would actually reduce the rate of aortic root dilatation in this population. We did not see a decrease in aortic root dilatation in our study, as has been reported in the mouse, but neither did we see an increase in diameter. Obviously, our statistical power is too low: it took more than 4 years and more than 1000 subjects to suggest that sartan might possibly lead to a reduction in the rate of aortic root dilatation^29^.

This benefit can be expected with a minimum of risk: during the CPET and the muscular exercise test (mET), the increase in blood pressure in MFS patients never exceeded 160 mmHg. It is important to note that, despite the limited increase in blood pressure, high heart rates were achieved during the training session, as shown in Table 3 (e.g. an average of 85% of peak HR). This suggests that vigorous intensity exercise can be proposed without risk. It is possible that the lower peak blood pressure observed during CPET at baseline (145±19.5 *vs.* 188±46.8; p<0.001) was related to the use of beta-blocker therapy in all patients.

In conclusion, MFS patients have altered QoL and exercise capacity, both of which can be improved by supervised home exercise training. We also show that many haemodynamic parameters are improved by training, which may translate into lower aortic stress and therefore limit aortic root dilatation, although we were not able to show such an effect in a short period of time in a limited population.

## Study limitations

The program used during the training session was individualised and close monitoring of heart rate and perceived exertion was carried out throughout the training period for all patients. This takes time and may limit the reproducibility of the results obtained here. However, a website is being set up to facilitate the adaptation of the sessions and to provide all the necessary information to the patients. The size of the population is limited, but the effect of training observed is impressive and unlikely to be the result of chance. Finally, the population included was selected to be low-risk and the applicability of the results obtained in higher-risk patients, who may benefit more from the training session, is unknown, particularly because of the haemodynamic risk.

### Non-standard abbreviations and acronyms

1RM: One repetition maximum
MFS: Marfan syndrome
FBN1: Fibrillin-1
MFS-C: Marfan syndrome control
MFS-T: Marfan syndrome training
H-S: Healthy subjects
FB: Fat body
MM: Mass muscular
BMI: Body mass index
EF: Ejection fraction
LV: Left ventricle
GLS: Global longitudinal strain
RV: Right ventricle
PWV: Pulse wave velocity
OUES: Oxygen uptake efficiency slope
VDO_2_peak: Peak oxygen uptake
VT1: First ventilatory threshold
VE: Minute ventilation
VDCO_2_: Volume of CO_2_ produced per minute
VE/VDCO_2_: Ventilation/carbon dioxide output ratio
FEV1: Forced expiratory volume in one second
mET: Muscular exercise testing
HR: Heart rate
SBP: Systolic blood pressure
DBP: Diastolic blood pressure
CPET: Cardiopulmonary exercise testing
Aix: Augmentation index
TGF-B: Transforming growth factor beta
TIMP: Tissue inhibitors of metalloproteinases
MMPs: Matrix metalloproteinases
QoL: Quality of life

## Data Availability

All data generated or analyzed during this study are included in this published article and its supplementary information files. Detailed data that support the findings of this study will be available.

## Acknowledgments

The authors dedicate this work to the patients who participated in the study and to all the healthcare professionals in the Marfan Syndrome Reference Center in Hopital Bichat. This study was presented at the European Days of the French Society of Cardiology in January 2022.

